# Evaluation of hemodynamic parameters in adult patients following administration of non-ionic intravenous contrast agents in computed tomography examinations in a tertiary hospital in Ghana

**DOI:** 10.1101/2024.11.23.24317583

**Authors:** R.N.A. Smillie, C. E. Kokah, G. Nunoo, A. Osei, B.O. Botwe, S. Anim-Sampong

**Affiliations:** Dept. of Radiography, University of Ghana School of Biomedical and Allied Health Sciences, Box KB 143, Korle Bu, Accra, Ghana; Dept. of Radiology, Korle Bu Teaching Hospital, Korle Bu, Accra, Ghana; Department of Diagnostics and Medical Imaging, University of Ghana Medical Centre, Legon, Accra, Ghana; Division of Midwifery and Radiography, School of Health & Psychological Sciences, City, University of London, Northampton Square, London EC1V 0HB, UK

**Keywords:** Computed tomography, contrast media, systolic blood pressure, diastolic blood pressure, heart rate

## Abstract

**Background:** Beside factors such as measurement technique, equipment accuracy, patient anxiety, race, ethnicity, ecological factors, diet (high sodium and calorie intake, and low potassium), physical inactivity, health conditions, and genetic vulnerability also influence blood pressure. Iodinated contrast agents (ICAs) or media (ICMs) are widely applied to improve the visibility of internal organs and other tissues in computed tomography (CT) and magnetic resonance imaging (MRI) procedures. ICAs have transient vasodilatory properties which can influence hemodynamic parameters immediately after administration during CT scans. Discrepancies on the effects of ICAs following their administration on hemodynamic parameters in adult patients who underwent contrast-enhanced CT (CECT) examinations have been reported in the literature. Anecdotal evidence further suggests that limited studies of the subject in Ghana. Knowledge of the relationship between hemodynamic parameters and contrast media is needed for prompt treatment, as well as the development of protocols to govern the administration of ICAs.

**Aim:** This study therefore evaluated HR, systolic blood pressure (SBP), and diastolic blood pressure (DBP) levels in adult patients following administration of non-ionic ICA during CECT examinations.

**Methodology:** Since data of the study variables were collected prospectively (before and after CT examinations) to determine SBP and DBP and HR levels following administration of non-ionic ICA during CT examinations, a prospective case-control study design was used, while a non-probability convenience sampling was employed to sample a population of 128 patients consisting of equal numbers of cases (experimental) and controls groups. Measurements of HR, SPB and DBP were made before and after the scans in both groups. Data analyses were done with Statistical Package for Social Sciences (SPSS) version 23. Pearson and Spearman correlations were used to compare the data variables obtained between the cases and control group. A *p*-value < 0.05 was considered statistically significant.

**Results:** The measured mean values of HR, SBP and DBP were higher after contrast administration (HR=84.75±14.00 bpm; SBP =128.39±17.98 mmHg; DBP= 80.00 ± 13.26 mmHg) than before (HR=82.56±15.08 bpm; SBP=120.81±14.32 mmHg; DBP=78.94±11.90 mmHg). There were insignificant differences between HR and SBP (*p* =0.716) and DBP (*p* = 0.533) prior to contrast media. The HR increase was statistically significant after contrast media (*p=*0.008). The mean differences in the HR, SBP, and DBP between genders were statistically insignificant after contrast administration.

**Conclusions:** Administration of non-ionic ICA increases HR but had no effects on SBP and DBP in both male and female adult patients who underwent diverse CECT examinations. The correlation statistics established no significant relationship between doses of contrast media and increases in the HR. No statistically significant differences between patient gender, BMI, and age on the hemodynamic parameters were found.

## Introduction

Iodinated contrast agent (ICAs) are used to opacify vascular structures, solid abdominal and pelvic organs, and widely applied in visualization of internal organs and other tissues through intravenous or intra-arterial, oral and rectal means, characterize soft tissue lesions, and to improve their diagnostic accuracies during non-invasive radiological procedures such as computed tomography (CT) and magnetic resonance imaging (MRI) procedures [1], [2]. The use of iodine in ICAs is attributed to its high contrast density, firm binding to the benzene molecule and low toxicity [3] X-ray energies are attenuated by covalently bonded iodine atoms because their atomic radius falls within the domain of x-ray wavelengths [4].Intravenously administered ICAs exhibit rheological, coagulatory, physiological, electrophysiological and hemodynamic effects owing to their viscosity, hydrophilicity, ionicity and contrast media pH [5]. Hence, despite the ability to enhance the visualization of anatomic parts, ICAs have the tendency to cause adverse effects. In particular, adverse effects of ICA including hypersensitivity reactions, thyroid dysfunction, and contrast-induced nephropathy, of which hypersensitivity reactions are the most common have been reported [6]. According to [1],contrast media administration is classified as the third most common cause of iatrogenic acute kidney injury, as well as higher chances of major adverse events such as initiation of dialysis, renal failure, stroke myocardial infarction and death.

Types of contrast media, consideration of severity of reaction and patients histories of allergies are factors which influence the ratings of anaphylactic reactions of radiographic ICAs whose transient vasodilatory properties influence hemodynamic parameters immediately after administration [7] Using iodixanol 320 and iomeprol 350 contrast media, [8] found no effects on tissue temperature, heart rate (HR), systolic blood pressure (SBP) and diastolic blood pressure (DBP), or cardiac output per minute in patients, while [9] reported insignificant effects of non-ionic low-osmolar contrast media (LOCM) on blood pressure in pheochromocytoma patients since there was no increase in plasma catecholamine levels. [10] also found no correlation between the mean doses of contrast material and its influence on hemodynamic parameters. On the contrary, recent studies have reported increases in HR, SBP and DBP following administration of non-ionic LOCM [11], [12].

According to [13]an estimated 50% of the approximately 76 million CT procedures performed annually use ICAs. Increasing numbers of CT radiological investigations imply increases in the adverse effects from ICAs and radiation-induced stochastic effects [14]. Despite the frequent use of contrast agents in CT and MRI examinations, there are very limited studies on the effects of administration of non-ionic contrast media on SBP, DBP and HRs in some geographical areas including Ghana. The lack of adequate knowledge on the effects of contrast media on patient hemodynamic parameters and HR limits the understanding of contrast effects on patients undergoing CT examination. Knowledge of the relationship between blood pressure and contrast media is needed for prompt treatment, as well as the development of protocols to govern the administration of ICAs. This study therefore evaluated the effects of intravascular administration of low-osmolar non-ionic contrast media on HR, SBP, and DBP before and after administration of non-ionic ICAs in contrast-enhanced CT (CECT) procedures in a tertiary hospital in Ghana. In particular, the study determined and compared the HRs, SBPs, and DBPs before and after the administration of ICAs, the influence of anthropometric factors on these hemodynamic parameters after the administration of contrast media, and also compared blood pressure values obtained after contrast injection with internationally accepted standards.

## Methods

This prospective case-control study was conducted at the radiology department of a tertiary referral hospital in Ghana from January to August 2019. Data on the study variables were collected prospectively (before and after CT examinations) to determine BP and HR levels following administration of non-ionic contrast media during CT examinations. Due to the absence of appointment system as well as the need for easy accessibility to patients at the hospital’s CT Unit, non-probability convenience sampling was used to sample a population of 128 patients (≧18 years) referred for CECT and non-CECT investigation for the study. Adult patients presenting with histories of hypertension or physiological conditions which predisposed them to hypertension were excluded. The study also excluded CECT examinations that exceeded 10 minutes after contrast administration.

A Cochran-determined sample size of 64 patients (cases) referred for CECT was obtained according to Eqn (1):

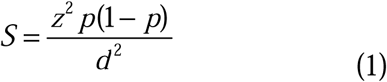

where *S* is the sample size, *z* is standard normal variate (=1.96 at 5% type 1 error (*p* <0.05), *d* is absolute error (0.05) and *p* (0.043) is expected proportion in population based on a previous study [15]. In order to ensure a precise data outcome, control data of 64 patients referred for CT examinations without contrast media were also collected.

All the 12 CT scans or procedures were performed on the 640-slice Toshiba Aquilion One CT scanner (Fig.1) where data acquisition was made from automatically generated volume-weighted CT dose index (CTDI_vol_) and dose length product (DLP) values for each patient scan based on radiographer selected parameters of the scan. Automatic dose reduction was also ensured by integration of Toshiba’s Adaptive Iterative Dose Reduction 3D (AIDR 3D) iterative reconstruction technology into the imaging chain [12].

**Figure 1:**
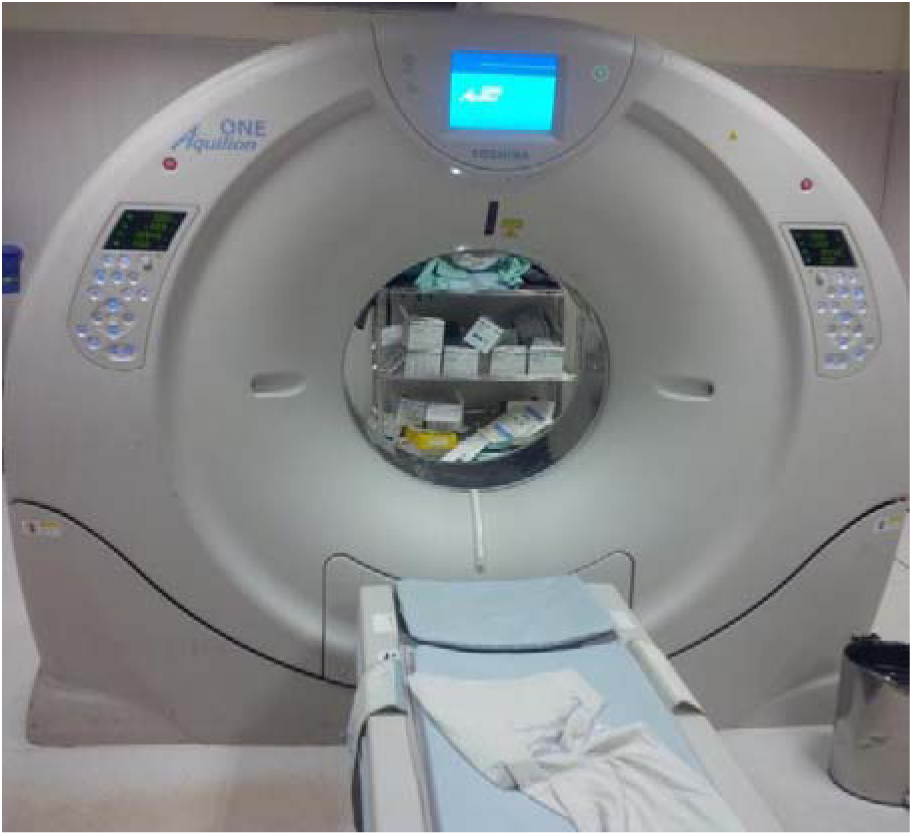
Toshiba Aquilion One CT Scanner.

Blood pressure (SBP and DPB) and HR were measured with a digital blood pressure monitor before and after the procedure in both cohorts of patient groups. To prevent errors, measurements were taken on the left arm of the patients seated with their arms supported against the chair, and elbows about heart level. The blood pressure cuff was placed around the unclothed and extended upper arm close to the heart around the brachial artery region after instructions pertaining to restricted movement and talking during measurements were given. The cuff was then inflated by clicking the power button on the blood pressure monitor.

For CECT of the abdominal regions, measurements prior to contrast examination were done with the patients seated, while the post-contrast administration measurements were performed during the delay phase with patients lying supine on the examination table. A 5-minute time gap between the two measurements for all the examinations was allowed. Comparison in values between the cases and the controls was done to estimate the effect of contrast media on blood pressure and pulse rate. Patient demographics [patient ID, age, gender, height, weight, body mass index (BMI)] and hemodynamic parametric data (SBP before (SBP-B), SBP after (SBP-A), DBP before (DBP-B), DBP after (DBP-A), HR before (HR-B), HR after (HR-A)) were recorded. Other data such as patient history, volume and type of contrast agent, region examined with or without contrast were also recorded.

Ethical approval and permission to conduct the study were granted by the Ethical and Protocol Review Committee (EPRC) of the University of Ghana School of Biomedical and Allied Health Sciences (SBAHS-RD/10557438/SA/2018-2019) and management of the Radiology Department of the hospital. Patient informed consent (written and verbal) was obtained from consenting patients whose identities were coded to ensure their anonymity and confidentiality. The process was clearly explained to them in their preferred languages of understanding. The patients also understood that they could opt out of the study for any reason at any point during the study.

### Data Analysis

The Statistical Package for Social Sciences (SPSS) version 23 was used for descriptive and inferential data analyses. Pearson and Spearman correlational statistics were performed to compare and establish associations and correlations between the experimental and control variable. A *p*-value < 0.05 was considered as statistically significant difference.

## Results

### Patient demographics

The population consisted of equal numbers (*n=*64, 50.0%) of male and female patients with mean ages of 47.2±16.1 years and 46.6±15.9 years respectively (Table 1). The population mean age was 46.92 ± 15.94 years (range: 19 to 87 years). The majority of patients were aged 30 – 39 years (*n=*31, 24.2%) and fewer patients 70 years and over (*n=1*2, 9.4%).

**Table 1:** Patient demographics: age, gender and BMI.

| Age group (yrs) | Frequency |  |  |  |  |  |
| --- | --- | --- | --- | --- | --- | --- |
|  | Male [*n* (%)] |  | Female [*n* (%)] |  | Total [*n* (%)] |  |
| 19 – 29 | 8 (6.2) |  | 13 (10.2) |  | 21 (16.4) |  |
| 30 – 39 | 16 (12.5) |  | 15 (11.7) |  | 31 (24.2) |  |
| 40 – 49 | 13 (10.2) |  | 7 (5.5) |  | 20 (15.6) |  |
| 50 – 59 | 8 (6.2) |  | 12 (9.4) |  | 20 (15.6) |  |
| 60 – 69 | 12 (9.4) |  | 12 (9.4) |  | 24 (18.8) |  |
| 70+ | 7 (5.5) |  | 5 (3.9) |  | 12 (9.4) |  |
| Total | 64 (50.0) |  | 64 (50.0) |  | 128 (100.0) |  |
| Mean age (yrs) | 47.23 ± 16.06 |  | 46.61 ± 15.94 |  | 46.92 ± 15.94 |  |
| Anthropometrics |  |  |  |  |  |  |
| Anthropometric factors | Mean±SD |  | Minimum |  | Maximum |  |
|  | M | F | M | F | M | F |
| Height (cm) | 171.78±7.78 | 164.88±6.86 | 140 | 145 | 186 | 183 |
| Weight (kg) | 67.55±11.38 | 71.30±18.23 | 35 | 37 | 94 | 135 |
| BMI (kg/cm<sup>2</sup>) | 22.87±3.49 | 27.27±7.27 | 15.62 | 14.82 | 29.06 | 53.75 |
| Contrast volume (ml) | 66.56±18.60 | 68.75±17.55 | 40 | 100 | 40 | 100 |

Key: SD=standard deviation, M= Male, F=Female

The male patients were generally taller (mean height= 171.78 ±7.78 cm; range:140 cm to 186 cm) than the female patients (mean height= 164.88 ± 6.86 cm; range=145 cm to 183 cm). On the contrary, the female patients were heavier (mean weight=71.30 ± 18.23 kg) and hence, had higher BMI (mean BMI= 27.27 ± 7.27 kg/cm^2^) than their male counterparts (mean weight=: 67.55 ±11.38 kg, mean BMI= 22.87 ±3.49 kg/cm^2^).

### Hemodynamic Parameters

Fig. 2 shows the measured hemodynamic parameters of the patients. Among the male patients, the highest mean values for pulse rates (HR-B=82.4 bpm), systolic (SBP=130.6 mmHg), and diastolic (DBP=81.3 mmHg) blood pressures were found in age groups 60-69 years, 50-59 years, and above 70-years. Similarly, for the female patients, the highest mean pule rate of 83.0 bpm was recorded among female patients aged 19-29 years and 30-39-years, while the highest mean systolic (SBP=129.3mmHg), and diastolic (DBP=82.0 mmHg) blood pressures were found in patients aged 40-49 years.

**Figure 2:**
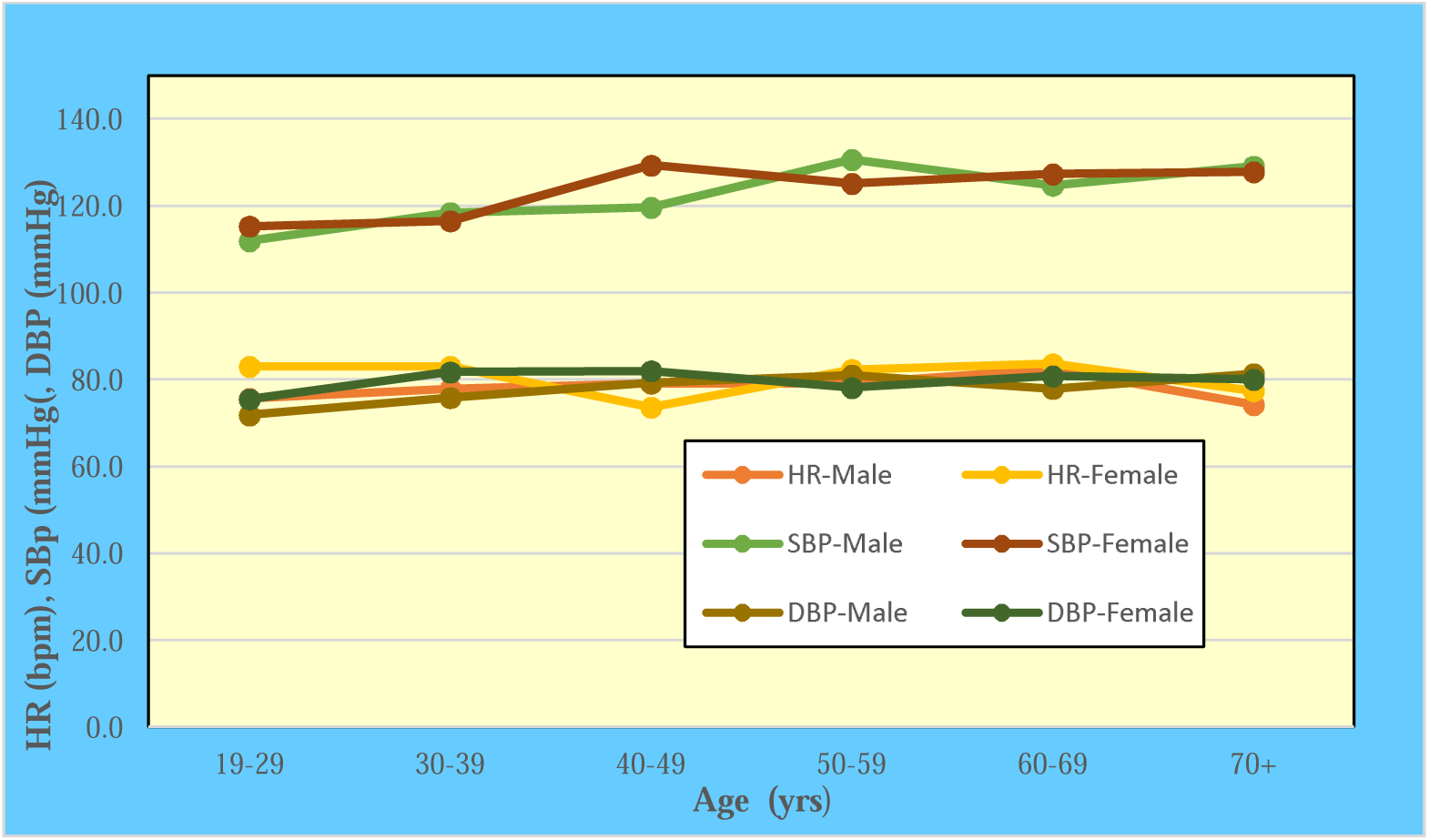
Hemodynamic parameters.

### Distribution of CT Examination by Body Parts

The types of CT procedures performed on the case and control patients is shown in Table 2.

**Table 2:** Distribution of CT examinations.

| Types of CT procedures | Cases group |  | Control group |  | Total |  |
| --- | --- | --- | --- | --- | --- | --- |
|  | Number | Percent, (%) | Number | Percent, (%) | Number | Percent, (%) |
| Abdominopelvic | 17 | 13.3 | 2 | 1.6 | 19 | 14.8 |
| Chest | 13 | 10.1 | 0 | 0.0 | 13 | 10.1 |
| Chest and abdomen | 4 | 3.1 | 0 | 0.0 | 4 | 3.1 |
| Cardiac angiogram | 1 | 0.8 | 0 | 0.0 | 1 | 0.8 |
| Cervical spine | 1 | 0.8 | 0 | 0.0 | 1 | 0.8 |
| Head | 23 | 18.0 | 58 | 45.3 | 81 | 63.3 |
| Head and neck | 2 | 1.6 | 0 | 0.0 | 2 | 1.6 |
| Neck | 1 | 1.6 | 1 | 0.8 | 2 | 1.6 |
| Neck and chest | 1 | 0.8 | 0 | 0.0 | 1 | 0.8 |
| Right Thigh | 1 | 0.8 | 0 | 0.0 | 1 | 0.8 |
| Shoulder | 0 | 0.0 | 2 | 1.6 | 2 | 1.6 |
| Thoracolumbar | 0 | 0.0 | 1 | 0.8 | 1 | 0.8 |
| Total | 64 | 50.0 | 64 | 50.0 | 128 | 100.0 |

Head (*n*=81 63.3%), abdominopelvic (*n=*19, 14.8%), and chest (*n*=13, 10.2 %) CT procedures were most requested. The majority of the head CT examinations (*n*=58, 45.3%) were performed among the control patients, while abdominopelvic (*n*=17, 13.3%) and chest (*n*=13, 10.3%) were mostly done among the case patients. Thoracolumbar, right thigh, neck and chest, cervical spine, and cardiac angiogram were least requested for both case and control groups.

### Contrast and Non-Contrast Enhanced CT Examinations

Contrast enhanced procedures were performed with either ultravist, omnipaque or iopamiro 370 non-ionic water-soluble ICA contrast media with pharmaco-kinetic characteristics of iopamidol 370 (Table 3). Out of the 64 CECT examinations, iopamiro ICA was used in 57 (89.1%) of the procedures. The volume (range: 40 ml to 100 ml) of injected ICA was based on the stature of the patients. Consequently, the mean contrast volumes administered to the female patients was marginally higher (68.75±17.55 ml) than in males (66.56 ±18.60 ml) due to their heavier weight and higher BMI as seen from Table 1.

**Table 3:** Pharmacokinetic characteristics of iopamidol.

| Concentration |  | Physical properties at 20<sup>0</sup>C |  |  | Osmometric values at 37°C |  |  |
| --- | --- | --- | --- | --- | --- | --- | --- |
| iodine/ml | iopamidol/100ml | Viscosity | Pressure | Relative | Osmolality | OP | pH |
| mg | g | (Nsm<sup>-2</sup>) | (mPa.s) | density | (osmol.kg<sup>1</sup>) | (Pa) |  |
| 150 | 30.6 | 0.3 | 1.5 | 1.17 | 0.342 | 8.7 | 7±0.5 |
| 200 | 40.8 | 3.3 | 2.0 | 1.22 | 0.413 | 10.5 | 7±0.5 |
| 300 | 61.2 | 8.8 | 4.7 | 1.33 | 0.616 | 15.7 | 7±0.5 |
| 370 | 75.5 | 20.9 | 9.4 | 1.41 | 0.796 | 20.3 | 7±0.5 |
| Types of administered ICAs in CECT examinations |  |  |  |  |  |  |  |
|  | Iopamidol | Omnipaque |  | Ultravist |  | Total |  |
| Number | 57 | 6 |  | 1 |  | 64 |  |
| Percent, % | 89.1 | 9.3 |  | 1.6 |  | 100.0 |  |
| Volumes of administered ICAs |  |  |  |  |  |  |  |
| Gender | Average volume, ml |  | Minimum, ml |  | Maximum, ml |  |  |
| Male | 66.56 ±18.60 ml |  | 40.00 |  | 40.00 |  |  |
| Female | 68.75±17.55 ml |  | 100.00 |  | 100.00 |  |  |

*OP=Osmotic pressure*

The relationship between patients’ weight and volume of ICAs is presented in Fig. 3.

**Figure 3:**
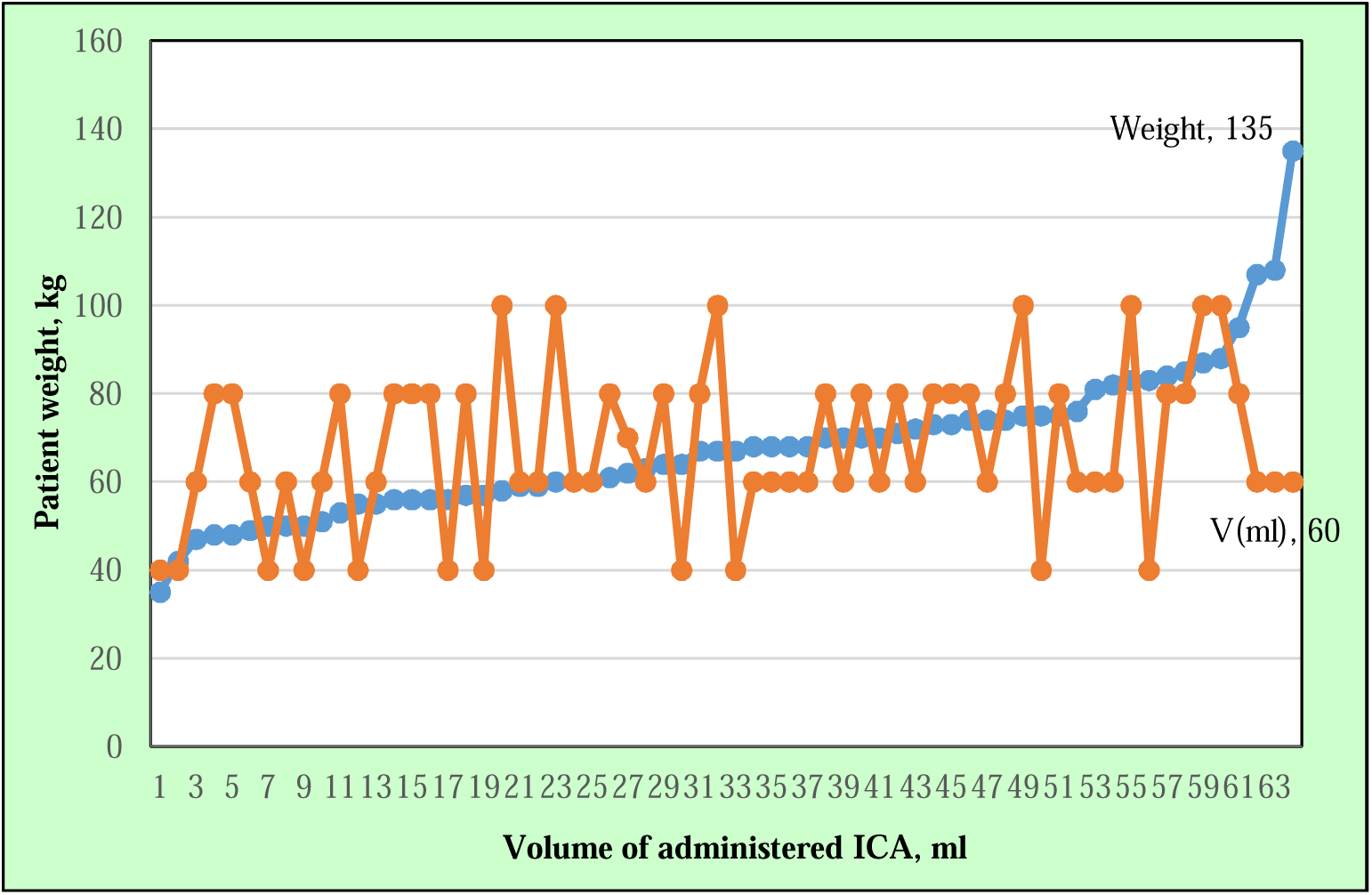
Relationship between weight and volume of contrast (*p=*0.115)

Patients with the minimum (*W*=35 kg) and maximum weights (*W*=135 kg) received 40ml and 60ml contrast media respectively. Patients who received the highest volume of contrast were mostly above 80 kg.

The distribution of patient indications for CECT examinations and patient history without contrast media are shown in Figs. 4 and 5 respectively. The majority of the patients referred for CECT examination presented with tumour (*n=*16, 25.0%), infection (*n*=16, 25.0%), and intracranial/space occupying lesion (IC/SOL) (*n=9*, 14.1%), while dyspnea, facial nerve palsy, deteriorating vision, ectopic kidney, hepatomegaly, bifascicular heart block, epilepsy were presented by 1 patient each. Headache (*n=*19, 29.7%), seizures (*n=*13, 20.3%), head injury (*n=12*, 18.8%), and CVA (*n=*6, 9.4%) were the most prevalent patient clinical histories.

**Figure 4:**
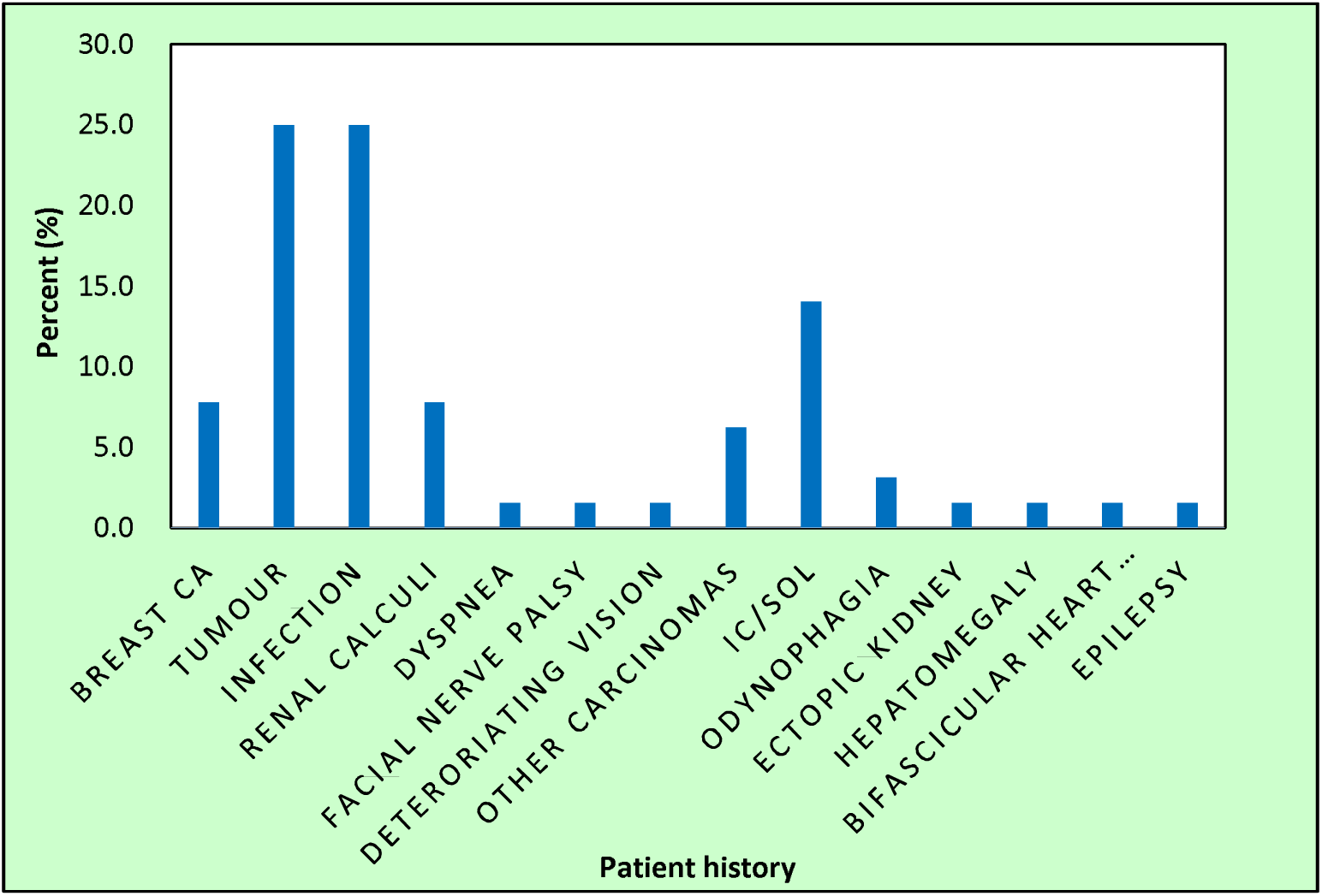
Patient indication for contrast enhanced examination.

**Figure 5:**
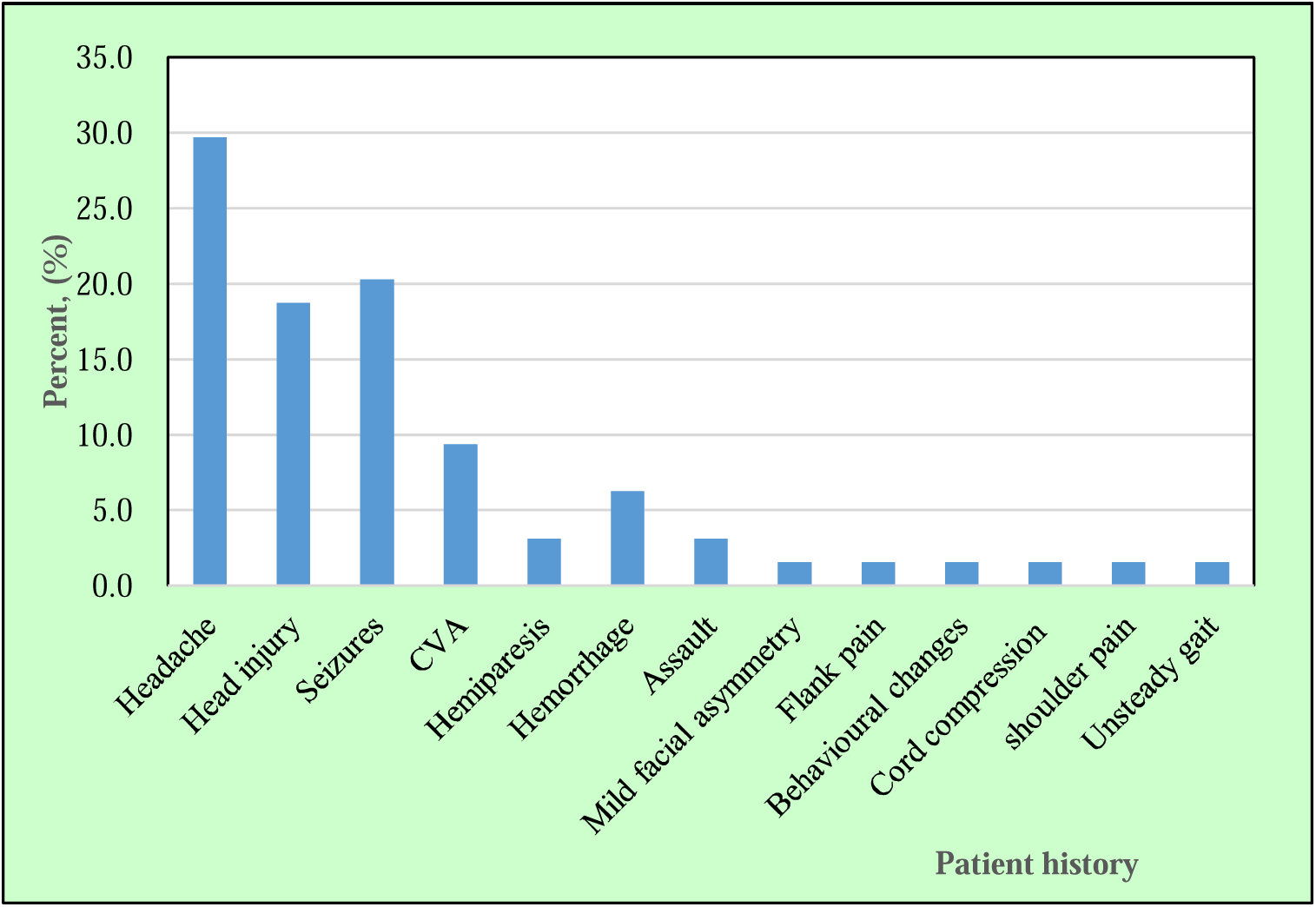
Distribution of patient history without contrast media.

### Inferential Statistics

The results of the comparison of hemodynamic values (case vrs. control groups) and effects of contrast media and demographics on blood pressure are presented in Tables 4 and 5 respectively.

**Table 4:**
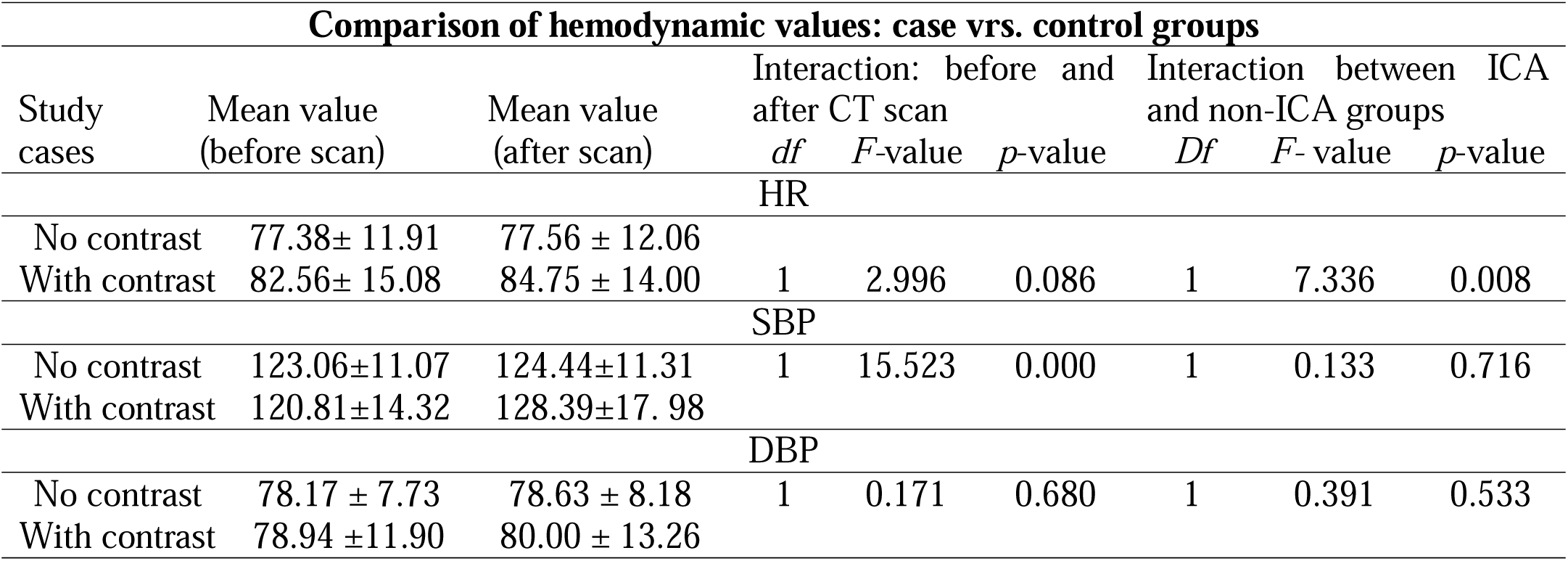
Comparison of hemodynamic parameters (case vrs. control groups) Comparison of hemodynamic values: case vrs. control groups.

| Comparison of hemodynamic values: case vrs. control groups |  |  |  |  |  |  |  |  |
| --- | --- | --- | --- | --- | --- | --- | --- | --- |
| Study cases | Mean value (before scan) | Mean value (after scan) | Interaction: before and after CT scan |  |  | Interaction between ICA and non-ICA groups |  |  |
|  |  |  | *df* | *F*-value | *p*-value | *Df* | *F*-value | *p*-value |
| HR |  |  |  |  |  |  |  |  |
| No contrast | 77.38± 11.91 | 77.56 ± 12.06 |  |  |  |  |  |  |
| With contrast | 82.56± 15.08 | 84.75 ± 14.00 | 1 | 2.996 | 0.086 | 1 | 7.336 | 0.008 |
| SBP |  |  |  |  |  |  |  |  |
| No contrast | 123.06±11.07 | 124.44±11.31 | 1 | 15.523 | 0.000 | 1 | 0.133 | 0.716 |
| With contrast | 120.81±14.32 | 128.39±17.98 |  |  |  |  |  |  |
| DBP |  |  |  |  |  |  |  |  |
| No contrast | 78.17 ± 7.73 | 78.63 ± 8.18 | 1 | 0.171 | 0.680 | 1 | 0.391 | 0.533 |
| With contrast | 78.94 ±11.90 | 80.00 ± 13.26 |  |  |  |  |  |  |

The measured mean HR, SBP and DBP with and without ICA administration in the case and control groups were higher after CT scan. The differences were however, higher among the ICA-administered case group (HR: 2.7%; SBP: 6.1%; DBP:1.3%) compared to the control group (HR: 0.2%; SBP: 1.1%; DBP:0.6%). Statistically, there was a significant effect of the ICA on HR (*p* = 0.008), but insignificant on SBP (*p* = 0.716) and DBP (*p =* 0.533). For heart rate (HR), the F-value for the interaction before and after the CT scan is 2.996 with 1 df and a p-value of 0.086, while the F-value for the interaction between ICA and non-ICA groups is 7.336 with 1 df and a p-value of 0.008, indicating a statistically significant difference in HR due to ICA administration. Conversely, for systolic blood pressure (SBP), a high F-value of 15.523 with 1 df and a p-value of 0.000 signifies a highly significant difference before and after the CT scan but no significant difference between ICA and non-ICA groups, where the F-value is 0.133 with 1 df and a p-value of 0.716. In the case of diastolic blood pressure (DBP), both interactions have low F-values (0.171 and 0.391) with 1 df and relatively high p-values (0.680 and 0.533), indicating no significant differences in DBP. The patients were age-categorized as young adults (19 – 39 yrs), middle aged adults (40 – 59 yrs) and old adults (≥ 60 yrs) (Table 5). Higher mean HR was measured for the middle aged adults (87.76±14.56 bpm) compared to the young (83.15±14.42 bpm) and old adults (84.35±13.22 bpm). The same was observed for the DBP. However, the mean SBP increased with age among the various age groups. No significant differences were found between the various age groups and effect of contrast media (*p* > 0.05). The differences in the mean HR between the male and female patients after contrast media administration were statistically insignificant (*p=*0.749). Similarly, there were no significant differences in the SBP (*p*=0.325) and DBP (*p=*0.975) in both female and male groups after administration of contrast media.

**Table 5:** Effects of contrast media and demographics.

| Comparison of effects of contrast media and gender on blood pressure |  |  |  |  |  |  |  |  |  |
| --- | --- | --- | --- | --- | --- | --- | --- | --- | --- |
| Demographic<br>s | HD<br>parameter | Mean<br>before<br>(mmHg) | Mean after<br>(mmHg) | Between time<br>point interaction<br>effects |  |  | Effect of<br>application and gender on<br>BP |  |  |
|  |  |  |  | df | F-value | p-value | df | F-value | p-value |
| Comparison of effects of contrast media between age group |  |  |  |  |  |  |  |  |  |
| Young | HR | 83.52±15.74 | 83.15±14.42 | 2 | 1.942 | 0.146 | 2 | 0.572 | 0.908 |
| Middle |  | 83.06±17.17 | 87.76±14.56 |  |  |  |  |  |  |
| Aged |  | 80.85±12.74 | 84.35±13.22 |  |  |  |  |  |  |
| Young | SBP | 113.59±13.91 | 120.30±14.73 | 2 | 0.174 | 0.840 | 2 | 0.944 | 0.392 |
| Middle |  | 125.41±13.21 | 133.88±15.58 |  |  |  |  |  |  |
| Aged |  | 126.65±11.83 | 134.65±20.22 |  |  |  |  |  |  |
| Young | DBP | 76.96±13.35 | 78.37±11.75 | 2 | 0.063 | 0.939 | 2 | 0.017 | 0.983 |
| Middle |  | 80.24±10.44 | 82.65±10.64 |  |  |  |  |  |  |
| Aged |  | 80.50±11.16 | 79.95±16.99 |  |  |  |  |  |  |
| Comparison of effects of contrast media between gender |  |  |  |  |  |  |  |  |  |
| Male | HR | 81.31±16.96 | 84.38±14.90 | 1 | 0.059 | 0.808 | 1 | 0.103 | 0.749 |
| Female |  | 83.81±13.09 | 85.13±13.27 |  |  |  |  |  |  |
| Male | SBP | 121.47±15.32 | 129.09±16.82 | 1 | 0.465 | 0.497 | 1 | 0.979 | 0.325 |
| Female |  | 120.16±13.47 | 127.69±19.31 |  |  |  |  |  |  |
| Male | DBP | 78.03±12.12 | 78.66±11.96 | 1 | 0.019 | 0.891 | 1 | 0.001 | 0.975 |
| Female |  | 79.48±9.17 | 81.34 ± 14.5 |  |  |  |  |  |  |
| Comparison of effect of contrast media between BMI |  |  |  |  |  |  |  |  |  |
| Underweight | HR | 80.92±12.98 | 86.08±11.55 | 3 | 2.881 | 0.039 | 3 | 2.579 | 0.057 |
| Normal weight |  | 87.19±16.54 | 85.85±15.64 |  |  |  |  |  |  |
| Overweight |  | 78.83±14.40 | 82.72±14.05 |  |  |  |  |  |  |
| Obese |  | 78.38±12.69 | 83.75±13.48 |  |  |  |  |  |  |
| Underweight | SBP | 111.08±16.14 | 118.17±20.12 | 3 | 0.735 | 0.533 | 3 | 0.918 | 0.435 |
| Normal weight |  | 118.88±12.90 | 126.96±13.22 |  |  |  |  |  |  |
| Overweight |  | 127.61±12.27 | 135.83±14.51 |  |  |  |  |  |  |
| Obese |  | 126.38±12.14 | 131.63±28.21 |  |  |  |  |  |  |
| Underweight | DBP | 73.67±15.17 | 73.50±13.98 | 3 | 0.375 | 0.771 | 3 | 0.071 | 0.975 |
| Normal weight |  | 78.46±12.35 | 82.00±13.35 |  |  |  |  |  |  |
| Overweight |  | 81.50±8.93 | 80.61±10.06 |  |  |  |  |  |  |
| Obese |  | 82.63±9.71 | 81.88±17.34 |  |  |  |  |  |  |

Table 5 further shows that the HR for underweight patients was highest among the four BMI categories (82.72 bpm), while the highest SBP and DBP were recorded among the overweight (135.83 mmHg) and normal weight (82.00 mmHg) patients respectively. There was no significant difference in relation to mean values obtained between all the hemodynamic parameters and the various categories of BMI after contrast media administration.

The Pearson and Spearman correlations between hemodynamic parameters and demographics (age, BMI) are shown in Table 6. The Pearson statistics showed positive correlations between age and SBP (*r* = 0.390, *p* = 0.001), and between age and DBP (*r=*0.086, *p=*0.499) after contrast. The relationships between age and HR as well as age and DBP were however, insignificant. Similarly, a statistically insignificant and negative correlation was identified between the contrast volume and DBP, while the positive correlations between contrast volume and HR (*r* = 0.020, *p* = 0.877) and SBP (*r*=0.152, *p=*0.231) were statistically insignificant.

**Table 6:** Pearson and Spearman correlations between hemodynamic parameters and demographics (age, BMI)

| Parameter | Pearson's<br>corr. (*r*) | *p*-value | Pearson's<br>corr. (*r*) | *p*-value | Pearson's<br>corr. (*r*) | *p*-value |
| --- | --- | --- | --- | --- | --- | --- |
|  | HR-A |  | SBP-A |  | DBP-A |  |
| Age (yrs) | -0.020 | 0.877 | 0.390 | 0.001 | 0.086 | 0.499 |
| Contrast volume | 0.020 | 0.877 | 0.152 | 0.231 | -0.010 | 0.938 |
| Parameter | Spearman's<br>corr. (*r<sub>s</sub>*) | *p*-value | Spearman's<br>corr. (*r<sub>s</sub>*) | *p*-value | Spearman's<br>corr. (*r<sub>s</sub>*) | *p*-value |
|  | HR-A |  | SBP-A |  | DBP-A |  |
| BMI (kg/cm<sup>2</sup>) | -0.041 | 0.747 | 0.352 | 0.004 | 0.189 | 0.134 |

*Key HR-A=HR after scan, SBP-A=SBP after scan, DBP-A= DBP after scan, corr.=correlation*

The HR and BMI correlated negatively (*r_s_*=-0.041) after contrast, while a positive correlation and statistically significant association between SBP and BMI (*r* = 0.352, *p*=0.004) was established. The association between BMI and DBP (*r* = 0.748, *p*=0.134) was insignificant.

## Discussions

### Demographic Characteristics

In case-control studies, it is necessary that every control group must consist of elements that exactly present the same features as the case group, except for applied interventions. In this regard, there were equal numbers of 64 patients in the case group who received ICA, and 64 patients in the control group who received no ICA. The wide age range of patients could be attributed to physician referrals of patients for other recommended imaging modalities other than CT in order to decrease the probability of carcinogenic risk following radiation exposure in the elderly.

### Contrast and non-contrast enhanced examinations

Head and spine were the most and least referred anatomic sites for both CECT and non-contrast CT examinations. Other studies have also reported head and spine CT examinations as the most common and least performed CT examinations [16]. Headache (29.7%) was the most prevalent indication presented by the patients. Consistent with this, [17] also reported headache as the most common indication (48 %) presented for CT scan. This could be explained by the fact that head CT scans are more often referred for adult patients who present with headache to rule out suspected tumor or subarachnoid hemorrhage since headache is a common symptom of brain tumor. Predominance of head CT scan over other body parts may be related to the consideration of CT and MRI as gold standards for identification of problems pertaining to the central and peripheral nervous system [18]. Also, CT scan is known for its ability to produce non-superimposed cross-sectional images of different anatomic parts such as bones, blood vessels and soft tissues inside the human body within the shortest period of time. This makes it possible for detection tiny fractures in head injuries and other abnormalities [19].

Most often, non-contrast head CT remains the primary technique for the initial evaluation of patients with suspected stroke [20] This is important because it excludes intracerebral hemorrhage and lesions that might mimic CVA, like a tumor [21]. A study has shown that the detection of hemorrhage via non-contrast CT has the advantage of fast acquisition and wide availability [22].This accounts for the higher numbers of non-contrast head exams compared to CECT for head exams as the ACR guidelines, suggest that non-contrast head CT is sufficient to diagnose abnormalities which would eliminate, the unnecessary administration of intravenous contrast media [23].

CECT was requested for the main indications (Fig. 4) because it is useful for staging and identification of tumour size. Iopamidol 370 LOCA was mostly used (89.1%) compared with iohexol and iopromide (1.56%) because of the higher adverse effects associated with iohexol (0.82%) and iopromide (0.65%) as compared with iopamidol (0.29%) reported by [24].

### Relationship between patients’ weight and volume of contrast

Body weight is the most important patient-related factor which impacts or influence the magnitude of vascular and parenchymal contrast media enhancement. Weight-based contrast administration protocols are therefore utilized in CECT modalities. This requires that higher contrast volumes are administered to heavier and larger patients due to their larger blood volumes. Consequently, administration of low volumes of ICA to heavy patients may result in image blurrness and compromised image quality. Repeat procedures resulting from poor or compromised image quality would further lead to increased patient radiation dose and elevated risk of biological damage, especially in the CT modality which is already associated with high patient radiation dose. It is therefore imperative that the correct volumes of ICAs are administered in CECT in order to avert these challenges.

In this study, the mean ICA volumes administered to female patients (68.75 ± 17.55 ml) was marginally higher because of their heavier weight and higher BMIs. [25] recommended administration of weight-related volumes of ICAs ranging from 30 ml (for pediatric patients), to 100 mls (for patients whose weight exceeded 120 kg). The lowest (40 ml for 35 kg patients) and highest (60 ml for 135 kg patients), as well as the mean contrast volumes applied in this study are within this range. The differences could be attributed to radiographers’ decisions regarding utilization of weight-based contrast administration protocols in CT imaging.

### Hemodynamic parameters and patient demographics

The study showed differences in the variation of HR with age among the male and female patients. The observed decreasing HR with age among females in this study is consistent with the literature where [26] reported similar findings. According to [27] the higher levels of estrogen in pre-menopausal women could account for this trend.

This present study showed increases in SBP and DBP with age up to 40-49 yrs for female patients, and 50 - 59 yrs in male patients (Fig.1). In a previous [28] reported similar findings of continuous increases in SBP between the ages of 30 and 84 years or over. The observed variations of SBP and DBP with age could be attributed to changes in arterial and arteriolar stiffness, and to increased peripheral vascular resistance in small vessels.

### Effect of contrast media on hemodynamic parameters

Post-contrast CT scans showed statistically significant increases in the patients’ HR. Other studies on post LOCM injection, and comparisons of the effects of iodixanol and ioxaglate during left ventriculography reported similar findings [12], [29]. The increases in HR could be attributed to the vasodilatory properties of contrast media and the effects of psychological factors resulting from heat sensation after contrast administration

Contrary to the findings of this study, [8] and [30] found no significant effects of iodixanol 320 and iomeprol 350 ICA on HR. According to [31] such inconsistencies could be explained by influences of physiochemical properties of contrast media, differences in volume of injected contrast, route of administration, radiographic procedures and study population on outcomes of hemodynamic parameters after contrast examination

This study found statistically significant differences in HR after contrast among patients who underwent CECT with non-ionic ICAs, while differences in the SBP and DBP were insignificant. The increase in SBP among the CECT patients (6.3%) was significantly higher compared to the almost negligible increase of SBP in control patients who received no contrast (0.3%). The increases in the DBP between the CECT and non-CECT examinations were also statistically insignificant. The findings of the current study are supported by [9] and [32] who reported no significant differences in terms of effects on arterial BP in in patients with coronary diseases after contrast administration or on LV pressures in patients undergoing contrast administration. In other studies, [8] found no effect of the contrast media on tissue temperature, SBP and DBP, or cardiac output per minute in patients who underwent CECT examination with iodixanol 320 and iomeprol 350, while [9] also reported insignificant effects of non-ionic LOCM on blood pressure in pheochromocytoma patients since there was no increase in plasma catecholamine levels.

On the contrary, [11]reported increases in SBP and DBP after administration of non-ionic ICA in patients, while [12] reported increases in SBP and DBP after using non-ionic (iohexol) contrast in assessing hemodynamic effects of contrast media in patients undergoing CECT examination. [33] reported a hypertensive crisis from a study in which non-LOCM caused an increase in pulmonary artery pressure during CT examination. In quantifying the effects on hemodynamic parameters and kidney function following intravenous administration of non-ionic LOCM with normal and low osmolality, [5] also observed a significant transient decrease in blood pressure following the administration of low-osmolar iopromide but not with iodixanol.

The observed discrepancies could be attributed to factors that influence outcome of hemodynamic parameters after contrast examination such as differences in volume of injected contrast, route of administration, physiochemical properties of contrast media, radiographic procedures and study population.

### Correlation between anthropometric factors and hemodynamic parameters

Post-contrast SBP correlated positively with age and BMI *(p=*0.001, *p=*0.004) (Table 6) as similarly reported [34], [35]. The absence of any effects of contrast volume on SBP and DBP is supported [10] who found no correlation between doses of contrast material and its influence on hemodynamic parameters.

This study found no statistically significant differences between patient demographics (gender, BMI, and age) in terms of measured blood pressure and HR. This is consistent with previous studies which found no difference between the incidence of adverse reactions to contrast media and patient characteristics such as sex, gender, weight, flow amount and flow ratio [10].

## Conclusion

The study found increases in HR but no effects of non-ionic ICA on SBP and DBP in both male and female adult patients who underwent diverse CECT examinations. Although no statistically significant differences between patient gender, BMI, and age on the hemodynamic parameters, an insignificant relationship was observed between the dose of contrast media and increase in the HR observed in this study. The measured SBP and DBP of the majority of patients in the cases were within the internationally recommended standards.

## Limitations

The high cost of CECT examinations limited the sample size as some patients who satisfied the inclusion criteria could be afford the fee. The effects of iopromide (ultravist) could not be compared with the other ICA since it was rarely used for scanning adult patients.

## Acknowledgement

The support of the radiology department of the hospital is duly acknowledged.

## Data availability

Data will be made available on request.

## References

[1] J. S. Hinson et al., “Risk of Acute Kidney Injury After Intravenous Contrast Media Administration,” Ann Emerg Med, vol. 69, no. 5, pp. 577–586.e4, May 2017, doi: 10.1016/j.annemergmed.2016.11.021.

[2] T. A. Rose and J. W. Choi, “Intravenous Imaging Contrast Media Complications: The Basics That Every Clinician Needs to Know,” Am J Med, vol. 128, no. 9, pp. 943–949, Sep. 2015, doi: 10.1016/j.amjmed.2015.02.018.

[3] M. V. Spampinato, A. Abid, and M. G. Matheus, “Current Radiographic Iodinated Contrast Agents,” Magn Reson Imaging Clin N Am, vol. 25, no. 4, pp. 697–704, Nov. 2017, doi: 10.1016/j.mric.2017.06.003.

[4] J. J. Pasternak and E. E. Williamson, “Clinical Pharmacology, Uses, and Adverse Reactions of Iodinated Contrast Agents: A Primer for the Non-radiologist,” Mayo Clin Proc, vol. 87, no. 4, pp. 390–402, Apr. 2012, doi: 10.1016/j.mayocp.2012.01.012.

[5] G. Widmann et al., “Systemic Hypotension Following Intravenous Administration of Nonionic Contrast Medium During Computed Tomography: Iopromide Versus Iodixanol,” Anesth Analg, vol. 126, no. 3, pp. 769–775, Mar. 2018, doi: 10.1213/ANE.0000000000002346.

[6] M. Andreucci, R. Solomon, and A. Tasanarong, “Side Effects of Radiographic Contrast Media: Pathogenesis, Risk Factors, and Prevention,” Biomed Res Int, vol. 2014, pp. 1–20, 2014, doi: 10.1155/2014/741018.

[7] S. KM. Ali, A. H. Stanford, P. J. McNamara, and S. Gupta, “Surfactant and neonatal hemodynamics during the postnatal transition,” Semin Fetal Neonatal Med, vol. 28, no. 6, p. 101498, Dec. 2023, doi: 10.1016/j.siny.2023.101498.

[8] K. Matschke, U. Gerk, C. Mrowietz, J.-W. Park, and F. Jung, “Influence of radiographic contrast media on myocardial oxygen tension: a randomized, NaCL-controlled comparative study of iodixanol versus iomeprol in pigs,” Acta radiol, vol. 48, no. 3, pp. 292–299, Apr. 2007, doi: 10.1080/02841850701209956.

[9] S. K. Baid, “Brief Communication: Radiographic Contrast Infusion and Catecholamine Release in Patients With Pheochromocytoma,” Ann Intern Med, vol. 150, no. 1, p. 27, Jan. 2009, doi: 10.7326/0003-4819-150-1-200901060-00006.

[10] X. Zhu et al., “Contrast material injection protocol with the flow rate adjusted to the heart rate for dual source CT coronary angiography,” Int J Cardiovasc Imaging, vol. 28, no. 6, pp. 1557–1565, Aug. 2012, doi: 10.1007/s10554-011-9950-y.

[11] A. M. John and S. Yadav, “EVALUATION OF BLOOD PRESSURE VARIATIONS DURING THE ADMINISTRATION OF INTRAVASCULAR CONTRAST MEDIA IN CECT ABDOMEN,” Asian Journal of Pharmaceutical and Clinical Research, vol. 11, no. 6, p. 309, Jun. 2018, doi: 10.22159/ajpcr.2018.v11i6.25284.

[12] S. Anim-Sampong, W. K. Antwi, B. Ohene-Botwe, and R. S. Boateng, “Comparison of 640-slice Aquilon ONE CT scanner’s measured dosimetric parameters with ICRP dose reference levels for head, chest and abdominal CT examinations,” Safety in Health, vol. 2, no. 1, p. 7, Dec. 2016, doi: 10.1186/s40886-016-0019-4.

[13] K. R. Beckett, A. K. Moriarity, and J. M. Langer, “Safe Use of Contrast Media: What the Radiologist Needs to Know,” RadioGraphics, vol. 35, no. 6, pp. 1738–1750, Oct. 2015, doi: 10.1148/rg.2015150033.

[14] D. Kobayashi, O. Takahashi, T. Ueda, G. A. Deshpande, H. Arioka, and T. Fukui, “Risk factors for adverse reactions from contrast agents for computed tomography,” BMC Med Inform Decis Mak, vol. 13, no. 1, p. 18, Dec. 2013, doi: 10.1186/1472-6947-13-18.

[15] U. Motosugi, T. Ichikawa, K. Sano, and H. Onishi, “Acute Adverse Reactions to Nonionic Iodinated Contrast Media for CT: Prospective Randomized Evaluation of the Effects of Dehydration, Oral Rehydration, and Patient Risk Factors,” American Journal of Roentgenology, vol. 207, no. 5, pp. 931–938, Nov. 2016, doi: 10.2214/AJR.16.16051.

[16] F. Zarb, L. Rainford, and M. McEntee, “Frequency of CT Examinations in Malta,” J Med Imaging Radiat Sci, vol. 42, no. 1, pp. 4–9, Mar. 2011, doi: 10.1016/j.jmir.2010.11.003.

[17] W. Shuaib, M. H. Tiwana, F. H. Chokshi, J. O. Johnson, H. Bedi, and F. Khosa, “Utility of CT head in the acute setting: value of contrast and non-contrast studies,” Irish Journal of Medical Science (1971 -), vol. 184, no. 3, pp. 631–635, Sep. 2015, doi: 10.1007/s11845-014-1191-3.

[18] Á. Bocskor, M. Hunyadi, and D. Vince, “National Academies of Sciences, Engineering, and Medicine (2015) The Integration of Immigrants into American Society. Washington, DC: The National Academies Press.,” Intersections, vol. 3, no. 3, Sep. 2017, doi: 10.17356/ieejsp.v3i3.401.

[19] G. Acharya, A. K. Mitra, and K. Cholkar, “Nanosystems for Diagnostic Imaging, Biodetectors, and Biosensors,” in Emerging Nanotechnologies for Diagnostics, Drug Delivery and Medical Devices, Elsevier, 2017, pp. 217–248. doi: 10.1016/B978-0-323-42978-8.00010-3.

[20] Md. D. Huda, “Characteristics of CT Findings in Patients of Acute Stroke and Their Relationship to Mortality,” 2018, Accessed: Nov. 18, 2024. [Online]. Available: http://rulrepository.ru.ac.bd/handle/123456789/73

[21] D. Birenbaum, L. W. Bancroft, and G. J. Felsberg, “Imaging in Acute Stroke,” Western Journal of Emergency Medicine, vol. 12, no. 1, p. 67, Feb. 2011, Accessed: Nov. 17, 2024. [Online]. Available: https://pmc.ncbi.nlm.nih.gov/articles/PMC3088377/

[22] M. P. Lin and D. S. Liebeskind, “Imaging of Ischemic Stroke,” CONTINUUM: Lifelong Learning in Neurology, vol. 22, no. 5, pp. 1399–1423, Oct. 2016, doi: 10.1212/CON.0000000000000376.

[23] A. C. Douglas et al., “ACR Appropriateness Criteria Headache,” Journal of the American College of Radiology, vol. 11, no. 7, pp. 657–667, Jul. 2014, doi: 10.1016/J.JACR.2014.03.024.

[24] S. Prakkamakul and S. Lerdlum, “Incidence and severity of acute adverse reactions to intravenous iodinated contrast media: 8-year experience in King Chulalongkorn Memorial Hospital,” Asian Biomedicine, vol. 7, no. 2, pp. 203–209, 2013, doi: 10.5372/1905-7415.0702.167.

[25] C. Saade, I. Alsheikh Deeb, M. Mohamad, H. Al-Mohiy, and F. El-Merhi, “Contrast medium administration and image acquisition parameters in renal CT angiography: what radiologists need to know,” Diagnostic and Interventional Radiology, vol. 22, no. 2, pp. 116–124, Jan. 2016, doi: 10.5152/dir.2015.15219.

[26] M. A. A. Santos, A. C. S. Sousa, F. P. Reis, T. R. Santos, S. O. Lima, and J. A. Barreto-Filho, “Does the Aging Process Significantly Modify the Mean Heart Rate?,” Arq Bras Cardiol, 2013, doi: 10.5935/abc.20130188.

[27] A. Voss, R. Schroeder, A. Heitmann, A. Peters, and S. Perz, “Short-Term Heart Rate Variability—Influence of Gender and Age in Healthy Subjects,” PLoS One, vol. 10, no. 3, p. e0118308, Mar. 2015, doi: 10.1371/journal.pone.0118308.

[28] E. Pinto, “Blood pressure and ageing,” Postgrad Med J, vol. 83, no. 976, pp. 109–114, Feb. 2007, doi: 10.1136/pgmj.2006.048371.

[29] B. Kojonazarov et al., “Evaluating Systolic and Diastolic Cardiac Function in Rodents Using Microscopic Computed Tomography,” Circ Cardiovasc Imaging, vol. 11, no. 12, Dec. 2018, doi: 10.1161/CIRCIMAGING.118.007653.

[30] J. Zhang et al., “Analysis of Heart Rate and Heart Rate Variation During Cardiac CT Examinations,” Acad Radiol, vol. 15, no. 1, pp. 40–48, Jan. 2008, doi: 10.1016/j.acra.2007.07.023.

[31] A. Caiazza, L. Russo, M. Sabbatini, and D. Russo, “Hemodynamic and Tubular Changes Induced by Contrast Media,” Biomed Res Int, vol. 2014, pp. 1–7, 2014, doi: 10.1155/2014/578974.

[32] R. Baglini, M. Sesana, C. Capuano, M. G. Rosella, C. Sardeo, and G. B. Danzi, “Left ventricular diastolic impairment during coronary arteriography with a non-ionic contrast medium.,” Minerva Cardioangiol, vol. 52, no. 4, pp. 323–8, Aug. 2004, [Online]. Available: http://www.ncbi.nlm.nih.gov/pubmed/15284681

[33] S. Nakano et al., “Hypertensive crisis due to contrast-enhanced computed tomography in a patient with malignant pheochromocytoma,” Jpn J Radiol, vol. 29, no. 6, pp. 449–451, Jul. 2011, doi: 10.1007/S11604-011-0573-Y/METRICS.

[34] N. K. Mungreiphy, S. Kapoor, and R. Sinha, “Association between BMI, Blood Pressure, and Age: Study among Tangkhul Naga Tribal Males of Northeast India,” Journal of Anthropology, vol. 2011, pp. 1–6, Dec. 2011, doi: 10.1155/2011/748147.

[35] M. Hosseini et al., “Blood pressure percentiles by age and body mass index for adults,” EXCLI J, vol. 14, pp. 465–477, 2015, doi: 10.17179/excli2014-635.

